# Child disciplining practices and their association with child stunting in Rwanda: insights from a population-based study

**DOI:** 10.64898/2026.08.11.26360229

**Authors:** Jean Nepo Utumatwishima, Ingrid Mogren, Kristina Elfving, Aline Umubyeyi, Gunilla Krantz

## Abstract

This study investigated the prevalence of physical and non-physical disciplining methods used by mothers of young children in Rwanda and examined their association with child stunting. The role of disciplining practices in child linear growth remains largely overlooked in interventions across low- and middle-income countries, including Rwanda, despite growing attention to psychosocial factors.

This cross-sectional study, conducted between November 15 and December 31, 2021, included 601 mother–child pairs selected through systematic random sampling. Child disciplining practices were assessed using the UNICEF Multiple Indicator Cluster Surveys questionnaire. Stunting, defined as chronic undernutrition during critical growth periods that leads to impaired growth and development, was measured using WHO standards (height-for-age Z score <−2 SD). Child disciplining methods were categorized into two types: physical (e.g., hitting or spanking) and non-physical (e.g., shouting or yelling). Multivariable logistic regression assessed the association between disciplining methods and stunting, adjusting for socioeconomic factors, IPV, and social support. The study included predominantly low-income, married mothers, >30 years old, with low educational attainment, and working in unskilled jobs as defined by the national occupational classification. Physical disciplining methods were used by 76.5% (*n*=449) of mothers, while 38.3% (*n*=225) used non-physical methods. Younger mothers used physical methods more than older ones (*p*=0.002). Mothers exposed to physical and sexual intimate partner violence (IPV) had a higher likelihood of using physical methods (*p*<0.001 and *p*=0.036, respectively). Exposure to psychological IPV was significantly associated with both non-physical (*p*=0.025) and physical methods (*p*=0.004). Of the 601 children, 27.1% (*n*=163) were stunted; 88.3% (*n*=144) were >1 year old. The odds of stunting were higher in older children subjected to both physical and non-physical methods (odds ratio [OR], 1.92; 95% confidence interval, 1.08–3.41). Using both disciplining methods was most strongly associated with stunting, with adjusted ORs ranging from 1.96 to 2.04.

Findings suggest disciplining methods may contribute to the risk of stunting, especially in older children in Rwanda. These findings highlight the urgent need for positive parenting education, child abuse prevention, and the integration of child protection screening into routine healthcare services in Rwanda. **Keywords** Child stunting, Child disciplining practices, Rwanda, Child abuse

## Introduction

For decades, physical punishment of children has been prevalent among parents as a disciplinary method, and it remains a global issue [1]. However, a fundamental shift in societal attitudes towards children has led to a global movement to recognize it as a form of child abuse [2]. Despite this global recognition, in Rwanda, discussions and interventions addressing child malnutrition and stunting have predominantly focused on socio-economic determinants such as poverty, food security, and access to healthcare [3–7]. The potential role of child abuse particularly violent maternal disciplining practices as a contributing psychosocial factor in child stunting remains largely overlooked [8]. While socio-economic factors have been extensively studied, psychosocial determinants such as maternal disciplining practices are consistently understudied, despite their plausible significance [9, 10]. This study provides the first empirical evidence in Rwanda demonstrating an association between abusive maternal disciplining practices and child stunting. Understanding these psychosocial influences is essential to developing more comprehensive and effective public health strategies aimed at combating child stunting, especially in rural areas where stunting rates remain high and the burden of child abuse may be underrecognized.

The United Nations Convention on the Rights of the Child forbids all types of violence against children [11]. Some countries in Europe, such as Sweden, introduced a ban on disciplinary violence against children in 1979, prohibiting all forms of violence and maltreatment of children, including corporal and mental punishment [12]. Child maltreatment is both prevalent and often concealed in clinical practice worldwide [13]. Only a small number of children disclose their experiences due to fear, confusion, denial, or a desire to protect the abuser, and even fewer receive the necessary support and assistance [14]. Child abuse and neglect have been central topics in scientific research for decades [15, 16]. There are four major types of child maltreatment: physical abuse, neglect, sexual abuse, and emotional abuse [17] . Physical abuse is the deliberate infliction of harm on a child, including hitting, kicking, biting, choking, burning, or causing other injuries [18–20]. Neglect is the failure to meet a child’s basic physical, medical, educational or emotional needs, including lack of care, supervision or support [18, 20, 21].

Emotional or psychological abuse refers to a repeated pattern of behaviour that undermines a child’s emotional growth or self-esteem [18, 20]. Childhood malnutrition and maltreatment, including abuse and neglect, remain significant public health challenges in low- and middle- income countries, worsened by poverty, food insecurity, and weak healthcare systems [22, 23]. In developed countries and those experiencing reductions in poverty, the incidence of child violence tends to decrease, which in turn can lead to improvements in child nutrition [24–26]. Indicators of concurrent malnutrition and maltreatment include not only stunted growth in both height and weight but also clinical signs, imaging results, and biological evidence, reflecting the complex nature of diagnosing abusive malnutrition [27]. Child abuse may contribute to child stunting through several interconnected mechanisms. First, psychobiological pathways linked to toxic stress play an important role. Repeated exposure to harsh or abusive discipline chronically activates the child’s stress response system, leading to elevated cortisol levels that can inhibit the secretion of growth hormones and impair normal physical development [28]. In addition, prolonged “fight-or-flight” states consume significant metabolic energy, forcing the body especially in food-insecure settings to prioritize immediate survival over growth in height and weight [29]. Second, nutritional neglect may occur in abusive environments. Excessive or punitive discipline may involve withholding food, while emotional trauma can reduce appetite or disrupt digestion, and chronic stress may cause gastrointestinal problems that limit nutrient absorption [30].

In low- and middle-income countries, child abuse often arises from excessive child disciplining [23, 31] whereby violent disciplinary practices towards children are a pervasive issue, particularly in sub-Saharan Africa [23, 31–34]. A study from Uganda, for example, reports that caregivers predominantly use harsh physical disciplinary methods, and women exhibit greater use of these practices compared with men [35]. Research has consistently demonstrated the association between child abuse and stunting, particularly in low- and middle-income countries [23, 33, 36, 37]. In Rwanda, the conversation surrounding child abuse predominantly centres on school-aged children and adolescents, which might neglect the experiences of younger children [38]. Notably, a study in 2022 revealed that an alarming 60% of children in Rwanda endure physical violence at some point between birth and the age of 18 years [38]. In addition, a 2015 survey of individuals aged 13–24 years found that 37% of young women and 60% of young men aged 18–24 years experience physical violence before reaching 18 years of age [39–41].

Furthermore, one study showed that medical doctors in Rwanda are not adequately trained to recognize and report child abuse in clinical practice [42]. The study revealed that only 16% of physicians had reported a case of abuse in the year preceding the survey; 38% had previously suspected abuse but chose not to report it [42]. This study examines disciplining practices used by mothers and their association with stunting in children aged between 1 month and 3 years, providing critical evidence to inform child protection policies, guide integrated intervention design, and strengthen public health strategies that address both psychosocial and nutritional factors affecting child growth in Rwanda.

## Methods

### Study design, study population and sample size

This cross-sectional study used interview-guided questionnaires to survey 601 mothers and their 601 children aged between 1 month and 3 years, residing in the Northern Province of Rwanda.

### Sample size calculation

The sample size for this study was determined on the basis of the estimated prevalence of child stunting in Rwanda, reported as 38% (43). To achieve a statistically robust estimate, we aimed for precision of ±4% with a 95% confidence level (*Z*=1.96). The calculated sample size was 630 after accounting for 10% non-responses as commonly observed in various health surveys and population studies.

### Sampling process

The sampling process followed a multistage random selection procedure, beginning with the random selection of 186 out of 2,743 villages in Rwanda’s Northern Province. The 186 villages were distributed across all districts of the Northern Province, namely Burera, Gakenke, Gicumbi, Musanze, and Rulindo. A geospatial grid system was applied by (1) overlaying a uniform grid across the province, (2) assigning each village to a corresponding grid cell, (3) using a random number generator to select grid cells and villages within them, and (4) ensuring spatially balanced representation of the selected areas. Within each selected village, community health workers identified households with mothers who had children aged between 1 month and 3 years for interviews. If a mother was unavailable in the initially selected household, the nearest household with an eligible mother was approached. Neighbouring households were chosen based on the assumption of similar sociodemographic characteristics, and this predefined procedure ensured that replacements were consistent across clusters, minimizing potential selection bias. Data collection faced limitations; weight and height measurements were not obtained from 29 households due to reasons such as congenital malformations in children or severe illness. Data from these households were excluded from the statistical analysis. This process resulted in an overall response rate of 95.4%, yielding a final sample of 601 mother–child dyads.

### Data collection procedures

We developed a culturally adapted survey that covered a range of factors to better understand the complex factors contributing to child stunting (S2 File. Questionnaire). The questionnaire was first translated into Kinyarwanda by certified linguists to maintain contextual accuracy and then reviewed by bilingual public health practitioners to ensure cultural relevance. It was subsequently pilot-tested with 35 mother-child pairs in Rwanda’s Northern Province, allowing us to refine wording and structure for improved clarity and reliability before large-scale deployment.

It included questions on how parents discipline their children (both physical and non-physical methods), household socio-economic conditions such as sanitation, mothers’ exposure to intimate partner violence (IPV) and the role of household support networks. To ensure accuracy and reliability, we followed World Health Organization (WHO) protocols to measure children’s height, length and weight.

The questionnaire was integrated into an Android-based application using the emGeo platform to enhance data management and efficiency. The study was led by the University of Rwanda’s School of Public Health, with a team of 13 experienced interviewers, including healthcare professionals and research trainees carrying out the fieldwork under the supervision of both Rwandan and Swedish researchers.

Before data collection began, the interviewers underwent rigorous training, which included a 5-day instructional phase followed by a 2-day field pilot in a village in the Northern Province. The data were collected over a 6-week period from November 15 to December 31, 2021.

### Measurement of dependent variables

The primary dependent variable in this study is child stunting, defined as a height-for-age Z score <−2. This threshold indicates that a child’s height-for-age is more than two standard deviations below the median of the WHO growth standards for their age and sex, indicating chronic undernutrition. Stunting, as a key measure of growth impairment, is calculated by measuring each child’s height and comparing it with age- and sex-specific WHO growth references, which provide a standardized population benchmark [44]. Anthropometric data, including weight and height, were collected following standardized WHO procedures. For children <2 years, recumbent length was measured using a ShorrBoard, and for children ≥2 years, standing height was measured. Weight was recorded using a SECA 874 digital scale. All measurements were taken twice, and the average value was used for analysis. In cases where the two measurements differed by >0.5 cm for height/length or >0.1 kg for weight, a third measurement was taken, and the median value was recorded. Data collectors were trained to ensure consistency, and inter-rater reliability was assessed during the training period [45].

### Measurement of independent variables

Socio-demographic factors were categorized and analysed as independent risk factors. The ages of the women and their partners were collected as continuous variables and categorized into 19–30 years and ≥30 years. Ages at marriage or cohabitation were collected as a continuous variable. In this study, age was dichotomized as a categorical variable as <18 years and ≥18 years. The number of children in the household was collected as a continuous variable and dichotomized as ≤3 children and >3 children. Marital status was categorized into five categories: cohabitating, divorced or separated, married, single mothers, and widowed, and further dichotomized into married or cohabitating and living without a partner (being single, divorced, widowed or separated). Educational levels for both mothers and their partners were reported in the following categories: never attended school, incomplete primary school, completed primary school, and secondary school or higher. These categories were then consolidated into two groups: (i) never attended school and primary level education, and (ii) secondary level education or higher. Occupations for both mothers and their partners were categorized into two groups: skilled workers and non-skilled workers. Child age was reported initially as a continuous variable and subsequently dichotomized into two groups: ≤12 months and >12 months. Similarly, family size was collected as a continuous variable and later categorized into families with ≤3 children and those with >3 children.

In addition to other socio-demographic variables, two key factors were assessed to evaluate the living conditions of the mothers: the availability of social support at the household level and exposure to IPV. Our previous research has shown that the absence of social support and IPV are associated with child stunting [4, 5].

The socio-demographic data used in this study are the same as those used in our previous studies. The Social Support Questionnaire-Short Form, which includes six items, was used to evaluate levels of social support [46]. Each question offered response options of never, sometimes, often and always. These responses were dichotomized into yes (i.e. sometimes, often, and always) and no (i.e. never). Social support was defined as the presence of a friend or family member who could provide assistance during illness, share food in moments of food insecurity or housing, lend money or offer psychological support in times of difficulty. Each item was converted into a yes or no response, with a positive response indicating available support used as the reference category, and a negative response placing the individual in the exposed category.

IPV before the pregnancy of the index child was assessed by measuring exposure across three categories using the WHO questionnaire: physical violence (six items), sexual violence (three items), and psychological abuse (four items) [47].

### Assessment of mothers’ use of child disciplining methods

The questionnaire on child discipline, supported by the United Nations International Children’s Emergency Fund (UNICEF) through the Multiple Indicator Cluster Surveys (MICS), was used to assess mothers’ use of child disciplinary methods. This tool was used to gather detailed information on mothers’ use of various disciplining practices, providing insights into their attitudes towards child discipline and potential child abuse [48]. The MICS questionnaire included a variety of response options to assess disciplining methods. Non-physical disciplinary methods included actions such as taking away privileges, forbidding activities the child enjoyed, restricting their movement outside the house, explaining why the behaviour was wrong, or suggesting alternative activities. Violent physical methods included shaking the child, shouting or yelling, spanking or hitting on the bottom with a bare hand, or using objects such as a belt, hairbrush, or stick to hit them on the bottom or other parts of the body. Additional responses involved verbal abuse, such as calling the child names such as dumb or lazy, and physical punishment targeting the face, head, ears, hands, arms or legs. More severe forms of physical punishment included repeated hitting or beating. Mothers were asked whether they had used any of the disciplining methods for the index child. Their response options were: Yes, I used this method or No, I did not use this method.

### Statistical analysis

The prevalence and frequency of mothers’ use of non-physical and/or physical child disciplining methods were estimated using counts (*n*) and percentages (%). Analyses were conducted using chi-squared tests for categorical associations between disciplining practices (physical, non-physical, or both) and child stunting, with statistical significance defined as *p*<0.05.

A composite variable for physical punishment was created by categorizing mothers who reported using any physical disciplinary methods. Similarly, a variable for non-physical punishment was constructed by grouping mothers who used non-physical methods. A variable for combined physical and non-physical punishment was created for mothers who used both approaches.

The association between mothers’ disciplinary practices and child stunting was examined using multivariable logistic regression models. The general form of the model is:

log[*P*(*Y_ij_*=1)/1−*P*(*Y_i_*=1)]=*β*_0_+*β*_1_*X*_1*i*_+*β*_2_*X*_2*i*_+*β*_3_*X*_3*i*_+*β*_4_*X*_4*i*_

where *Y_i_* is the binary outcome variable for child *i* (1=stunted, 0=not stunted), *P*(*Y_i_*=1) represents the probability of stunting, *β*_0_ is the intercept, *X*_1*i*_ represents socio-demographic factors (maternal age, education, occupation) included in model 2, *X*_2*i*_ represents social support, added in model 3, *X*_3*i*_ represents IPV exposure, added in model 4, *X*_4*i*_ represents interaction terms between disciplinary methods, social support, and IPV, included in model 5, and *β_k_* are the regression coefficients estimating the independent effect of each covariate on the outcome. The modelling strategy involved incremental adjustment across five models (models 1–5) to estimate adjusted odds ratios (aORs) and 95% confidence intervals (CIs). Model 1 included unadjusted (crude) estimates. Model 2 adjusted for key socio-demographic factors, including mother’s age, age at marriage, occupation, education level, and marital status. Model 3 further adjusted for household-level social support variables. Model 4 incorporated mothers’ exposure to IPV, considering physical, sexual, and psychological forms. Finally, Model 5 included interaction terms between disciplining methods, social support, and IPV to examine potential moderating effects on child stunting. This approach allowed for multivariable adjustment to control for confounding factors while assessing the independent association of each predictor with child stunting.

All analyses were performed using Stata 18.0 (StataCorp LP), with missing data handled via listwise deletion which was deemed appropriate due to the relatively low proportion of missing values and the assumption that data were missing completely at random, minimizing potential bias in the estimates.

### Ethical considerations

The study was approved by the Institutional Review Board of the University of Rwanda, College of Medicine and Health Sciences (approval no. 181/CMHS IRB/2021) and by the Rwanda National Health Research Committee (protocol no. NHRC/2020/PROT/047), and informed consent was obtained from all participants. Participation in this study was voluntary for all invited mothers. Written informed consent was obtained from all participants. Consent for children ≤3 years of age born to the participating mothers was provided by their mothers. These children were assessed for their general health and growth status through anthropometric measurements. Participants were not compensated for their participation. Before the interview, each mother received a comprehensive explanation of the questionnaire, assurance of confidentiality, and the option to withdraw at any time without consequences, both during and after the interview. Only one mother per household was interviewed to ensure participant safety and privacy, and any partner-related information was provided directly by the mother. Interviews were conducted privately between the participant and interviewer. If interruptions occurred, interviewers were trained to either pause the interview or shift to less sensitive topics until privacy was re-established. In cases where immediate healthcare was needed, the study team arranged transportation to appropriate healthcare facilities.

## Results

### Socio-demographic characteristics of participants and the prevalence of child disciplining methods

The study included 601 women and their 601 children. Most women (88.4%, *n*=531) were married, and 94% (*n*=565) were married at ≥18 years of age. The age of the participants varied; the largest group (56.4%, *n*=339) were aged ≥30 years (Table 1). Most of the women (78.2%, *n*=470) had completed only primary or no formal education, and 91.7% (*n*=505) worked in non-skilled jobs. Both mothers and their partners predominantly came from low-income backgrounds, as reflected in their educational level and employment status. Most women had limited education; most had only completed primary schooling or had no formal education (Table 1). The prevalence of child disciplining methods varied based on the socio-demographic characteristics of the women and the age of the child. Physical punishment was more common among younger mothers compared with mothers >30 years (*p*=0.004) (Table 1). Regarding household characteristics, mothers without shelter support demonstrated a statistically significant higher likelihood of using non-physical punishment (*p*=0.023).

**Table 1.**
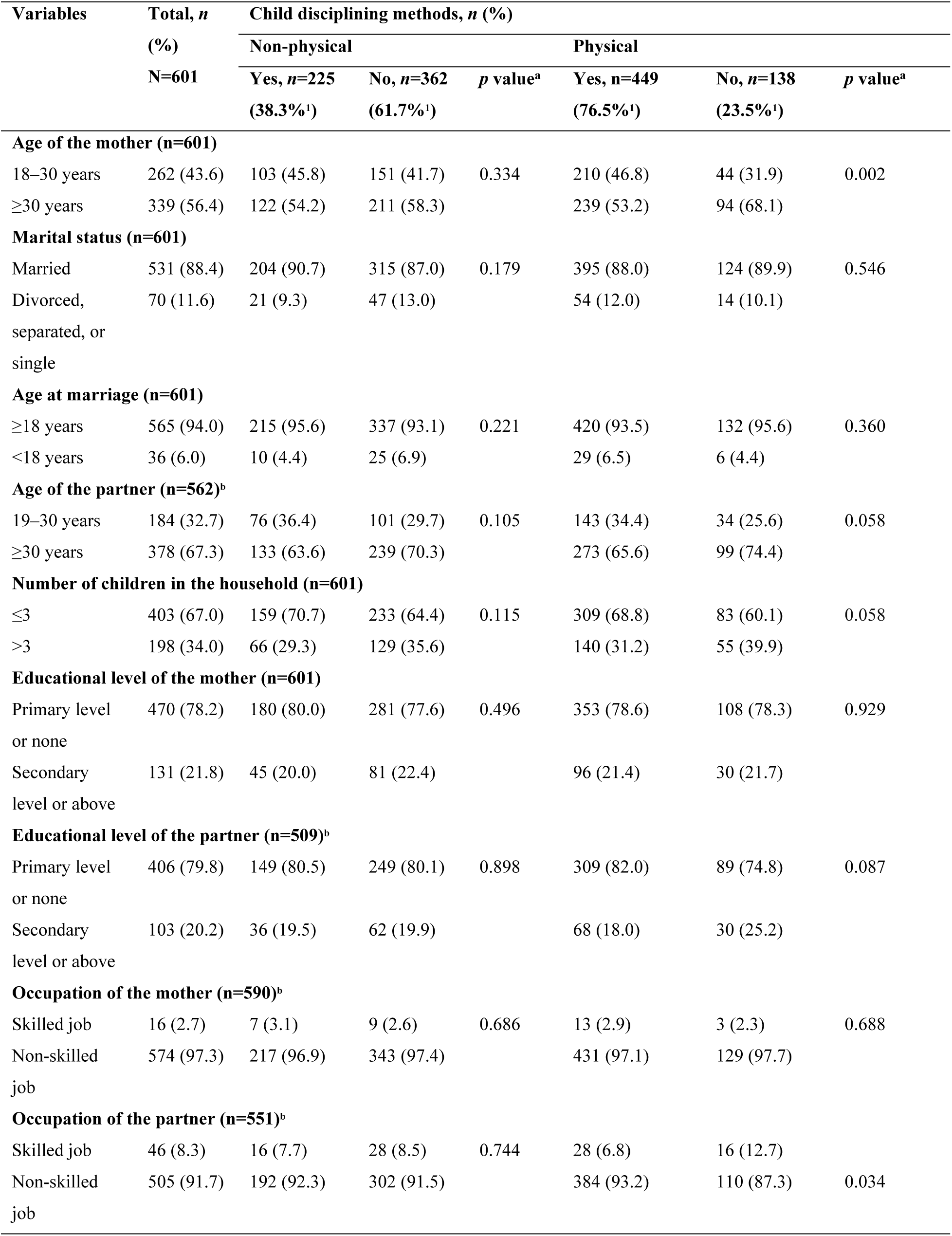
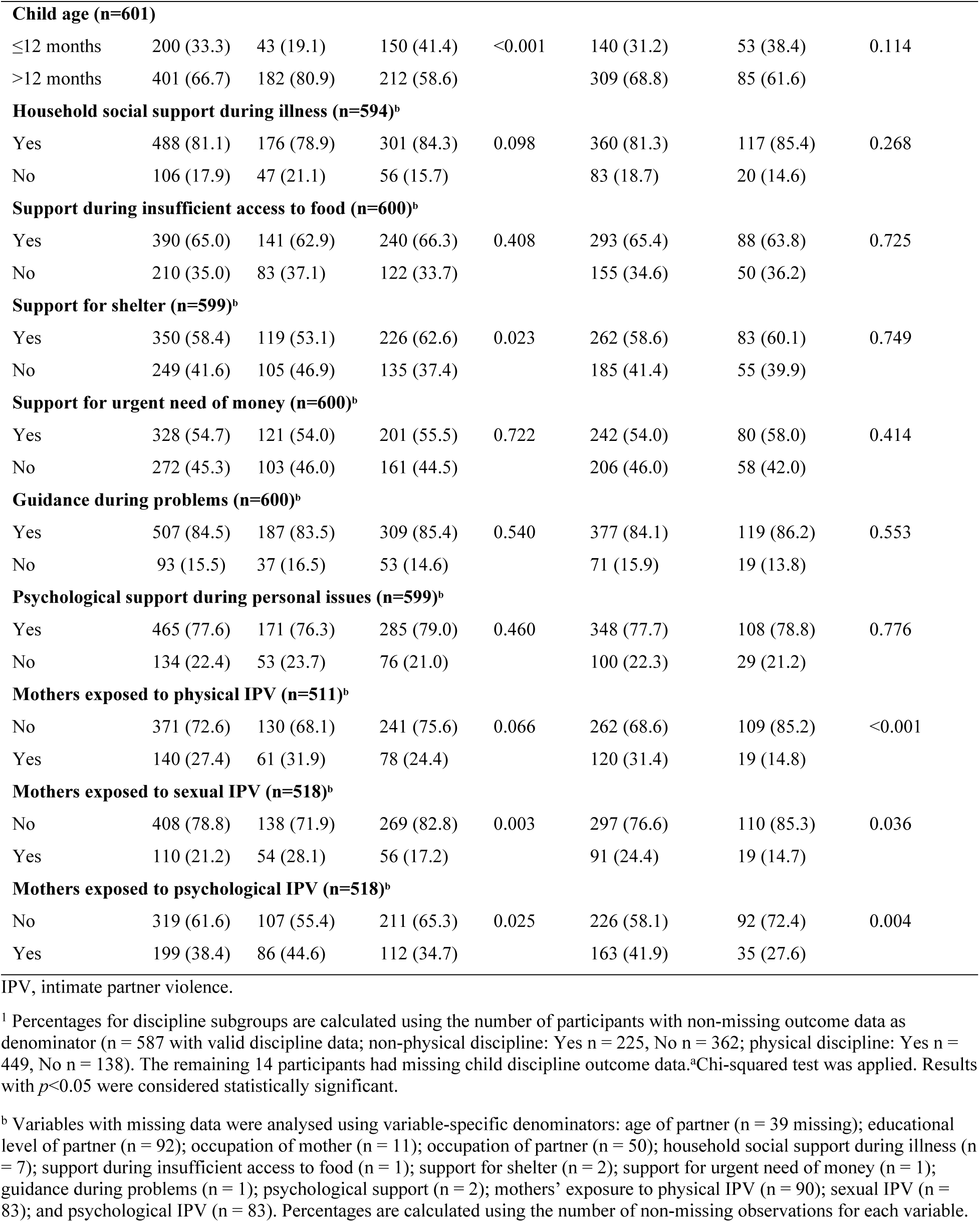
Socio-demographic characteristics of participants and prevalence of child disciplining methods with corresponding *p* values from chi-squared tests

Furthermore, mothers’ exposure to IPV played a significant role in the use of child disciplining methods. Mothers exposed to physical IPV were significantly more likely to use physical punishment (*p*<0.001), and those exposed to sexual IPV also showed a significant association with the use of physical punishment (*p*=0.036). In addition, mothers exposed to psychological IPV demonstrated a statistically significant association with the use of both non-physical and physical punishment (*p*=0.025 and *p*=0.004, respectively).

### Associations between child disciplining methods and child stunting stratified by the age of the child

Of the 601 children included in the study, 27.1% (*n*=163) were identified as stunted. When stratified by age, most (88.3%, n = 144) stunted children were in the group aged >12 months. Among the disciplining methods used by women, physical punishment was more commonly observed in children aged >12 months (Table 2). Regarding combined physical and non-physical punishment, 30.6% (*n*=141) of children aged ≤12 months and 69.4% (*n*=320) of children aged >12 months were subjected to both methods. Among older children, exposure to both physical and non-physical disciplining methods was significantly associated with an increased likelihood of stunting, with an odds ratio (OR) of 1.92 (95% CI, 1.08–3.41).

**Table 2.** Prevalences and associations between child disciplining methods (physical, non-physical, and combined) and child stunting stratified by age group (≤12 months and >12 months)

| Variables | Children's age |  |  |  |
| --- | --- | --- | --- | --- |
| | $\leq 12$ months | | $>12$ months | |
|  | No. | Percent | No. | Percent |
| Child stunting ($n=163$) | 19 | 11.7 | 144 | 88.3 |
| Physical discipline ($n=449$) | 140 | 31.2 | 309 | 68.8 |
| Physical discipline × child stunting | 15 | 78.9 | 117 | 83.6 |
| OR (95% CI)<sup>a</sup> | 1.47 (0.46–4.65) |  | 1.64 (0.97–2.79) |  |
| Non-physical discipline ($n=225$) | 43 | 19.1 | 182 | 80.9 |
| Non-physical discipline × child stunting | 3 | 15.8 | 72 | 51.4 |
| OR (95% CI)<sup>b</sup> | 0.63 (0.17–2.26) |  | 1.39 (0.92–2.09) |  |
| Both physical and non-physical discipline ($n=461$) | 141 | 30.6 | 320 | 69.4 |
| Child Stunting and both physical and non-physical discipline | 15 | 78.9 | 122 | 87.1 |
| OR (95% CI)<sup>c</sup> | 1.43 (0.45–4.52) |  | **1.92 (1.08–3.41)<sup>d</sup>** |  |

<sup>a</sup>Univariate regression analysis examining the association between physical discipline and child stunting
<sup>b</sup>Univariate regression analysis examining the association between non-physical discipline and child stunting
<sup>c</sup>Univariate regression analysis examining the association between combined physical and non-physical discipline and child stunting
<sup>d</sup>Bold denotes statistical significance

### Prevalence of child disciplining methods and their association with child stunting

Physical punishment methods were more prevalent; 76.5% (*n*=449) of mothers used such methods overall. Hitting on the bottom with hands (37.5%, *n*=220) was the most prevalent, followed by hitting with a belt (15.4%, *n*=90). Statistically significant associations with child stunting were observed for hitting on the bottom with hands, hitting with a belt, and the composite index for the use of physical punishment (*p* values of 0.006, 0.014 and 0.023, respectively) (Table 3). Non-physical punishment methods were less prevalent; 38.3% (*n*=225) of women used these methods overall. Shouting/yelling/screaming (29.7%, *n*=174) and privilege deprivation (30.3%, *n*=177) were the most frequently used non-physical methods. Among these, shouting/yelling/screaming, calling names, assigning another task, explaining wrong behaviour, and the composite index for non-physical punishment showed statistical significance for child stunting (Table 3).

**Table 3.** Association between child disciplining methods and child stunting: a stratified analysis of the use of physical and non-physical punishment

| Child disciplining methods | Mothers who used various child disciplining methods, *n* (%) | Child stunting, *n* (%) |  | *p* value<sup>a</sup> |
| --- | --- | --- | --- | --- |
|  |  | Yes, *n*=163 (27.1%) | No, *n*=438 (72.9%) |  |
| **Physical punishment** |  |  |  |  |
| Shaking (*n*=585) | 70 (12.0) | 25 (15.8) | 45 (10.5) | 0.080 |
| Hitting (bottom with hands) (*n*=586) | 220 (37.5) | 74 (46.5) | 146 (34.2) | 0.006 |
| Hitting (with belt) (*n*=585) | 90 (15.4) | 34 (21.4) | 56 (13.1) | 0.014 |
| Hitting (on hand/arm/leg) (*n*=207) | 31 (15.0) | 8 (17.4) | 23 (14.3) | 0.603 |
| Beating up (*n*=577) | 9 (1.6) | 4 (2.6) | 5 (1.2) | 0.219 |
| Aggregate Composite Index for physical punishment preference (*n*=587) | 449 (76.5) | 132 (83.0) | 317 (74.1) | 0.023 |
| **Non-physical punishment** |  |  |  |  |
| Privilege deprivation (*n*=585) | 177 (30.3) | 55 (34.8) | 122 (28.6) | 0.145 |
| Explaining wrong behaviour (*n*=208) | 89 (43.0) | 27 (57.4) | 62 (38.7) | 0.023 |
| Shouting/yelling/screaming (*n*=586) | 174 (29.7) | 62 (39.0) | 112 (26.2) | 0.003 |
| Calling names (dumb, lazy) (*n*=584) | 89 (15.2) | 33 (20.9) | 56 (13.1) | 0.021 |
| Giving another task (*n*=585) | 18 (3.1) | 9 (5.7) | 9 (2.1) | 0.026 |
| Aggregate Composite Index for psychological punishment preference (*n*=587) | 225 (38.3) | 75 (47.2) | 150 (35.1) | 0.007 |

<sup>a</sup>Chi-squared test was applied. Results with $p < 0.05$ were considered statistically significant.

### Association between disciplining methods and child stunting in multilevel regression analysis

The use of physical punishment was consistently associated with higher odds of child stunting across all models, with aORs ranging from 1.65 to 1.75 (Table 4). Similarly, the use of non-physical punishment was significantly linked to child stunting, with aORs between 1.61 and 1.74 (Table 4). Notably, mothers who used both physical and non-physical punishment exhibited the strongest association with child stunting, with aORs ranging from 1.96 to 2.04 across the models (Table 4). These findings remained significant after adjusting for socio-demographic factors, levels of social support, and mothers’ exposure to IPV, as well as their interactions.

**Table 4.** Mothers’ use of child disciplining methods and its association with child stunting, adjusted consecutively for socio-demographic variables, social support and intimate partner violence

| Types of disciplining methods used by mothers | Child stunting, OR (95% CI) |  |  |  |  |
| --- | --- | --- | --- | --- | --- |
|  | Model 1 | Model 2 | Model 3 | Model 4 | Model 5 |
| Physical punishment (n=449) | 1.71 (1.07–2.73) | 1.65 (1.02–2.68) | 1.68 (1.05–2.69) | 1.75 (1.09–2.79) | 1.71 (1.07–2.74) |
| Non-physical punishment (n=225) | 1.65 (1.14–2.39) | 1.74 (1.19–2.55) | 1.61 (1.11–2.34) | 1.68 (1.16–2.44) | 1.64 (1.13–2.38) |
| Use of both physical and non-physical punishments (n=461) | 1.99 (1.21–3.30) | 1.96 (1.16–3.29) | 1.96 (1.18–3.24) | 2.04 (1.23–3.39) | 2.00 (1.20–3.31) |

Model 1, crude (unadjusted) analysis; model 2, adjusted for socio-demographic factors, including mother's age, age at marriage, mother's occupation, education level and marital status; model 3, adjusted for socio-demographic factors and social support; model 4, adjusted for socio-demographic factors, social support and maternal exposure to intimate partner violence; model 5, includes interaction terms between disciplinary practices, social support and intimate partner violence.

### Analysis of child disciplining methods and their association with child stunting, considering categories of household social support and mothers’ exposure to intimate partner violence

The analysis revealed consistently higher ORs for child stunting across different child disciplining methods, irrespective of social support categories or types of IPV (Table 5).

**Table 5.** Analysis of child disciplining methods and their association with child stunting, considering categories of household social support and mothers’ exposure to intimate partner violence

| Child disciplining<br>method | Child stunting, adjusted odds ratio (95% confidence interval) |  |  |  |  |  |  |  |  |  |
| --- | --- | --- | --- | --- | --- | --- | --- | --- | --- | --- |
|  | Crude OR | Social support category |  |  |  |  |  | Type of intimate partner violence |  |  |
|  |  | Illness | Food | Shelter | Money | Personal<br>problem | Psycho-<br>logical<br>guidance | Physical | Sexual | Psycho-<br>logical |
| Physical punishment<br>preference (*n*=449) | 1.71<br>(1.07–2.73) | 1.67<br>(1.04–2.68) | 1.73<br>(0.91–1.94) | 1.70<br>(1.06–2.71) | 1.70<br>(1.06–2.72) | 1.70<br>(1.06–2.72) | 1.69<br>(1.06–2.71) | 1.57<br>(0.96–2.57) | 1.72<br>(1.06–2.79) | 1.78<br>(1.09–2.91) |
| Non-physical punishment<br>preference (*n*=225) | 1.65<br>(1.14–2.39) | 1.62<br>(1.12–2.36) | 1.65<br>(1.14–2.39) | 1.65<br>(1.14–2.39) | 1.66<br>(1.15–2.41) | 1.65<br>(1.14–2.40) | 1.65<br>(1.14–2.39) | 1.67<br>(0.69–1.80) | 1.71<br>(1.15–2.54) | 1.80<br>(1.21–2.67) |
| Preference of both<br>physical and non-physical<br>punishment (*n*=461) | 1.99<br>(1.21–3.30) | 1.94<br>(1.17–3.21) | 2.01<br>(0.90–3.32) | 1.99<br>(1.20–3.29) | 1.99<br>(0.85–3.29) | 2.01<br>(1.22–3.33) | 2.00<br>(1.21–3.30) | 1.90<br>(1.12–3.21) | 2.08<br>(1.24–3.49) | 2.19<br>(1.29–3.73) |

726 Crude OR: unadjusted odds ratios for the association between disciplinary preferences and child stunting.

Regarding the use of physical punishment, ORs ranged from 1.67 to 1.78; the highest association was observed among mothers exposed to psychological IPV (Table 5). The ORs for the use of non-physical punishment were between 1.62 and 1.80; the strongest link was seen in mothers exposed to sexual IPV. The use of both physical and non-physical punishment had the highest ORs, ranging from 1.90 to 2.19, particularly in contexts of exposure to psychological and sexual IPV. Among social support categories, consistent associations were observed; mothers lacking support for illness, shelter, and psychological guidance showed slightly higher ORs.

## Discussion

This study is among the first in Rwanda to examine mothers’ use of child disciplining methods and their associations with child growth outcomes, particularly stunting. Using multivariable regression models that adjusted for relevant maternal, child, and household characteristics, the study revealed significant associations between different child disciplining methods and higher odds of stunting among children in rural Rwanda. It also investigated the impact of household-level social support and women’s exposure to IPV on mothers’ use of child disciplining methods, which in turn significantly increased the risk of stunting in children.

### Maternal age and physical punishment of children

The analysis comparing non-physical and physical disciplinary measures showed that maternal age was associated with disciplinary approaches. Younger mothers were more likely to report the use of physical punishment than older mothers. Notably, mothers aged 18–30 years exhibited greater use of physical violence towards their children than mothers >30 years. Only mothers >30 years old demonstrated a reduced use of physical punishment, suggesting a significant shift in disciplinary practices with increasing parenting experience. The results of our study are in agreement with other studies where it has been shown that parental age influences the use of physical punishment, with younger parents tending to use physical punishment more frequently [49–51]. In Rwanda, the use of physical violence towards children among mothers, particularly those aged 18–30 years, may be linked to limited parenting experience and own exposure to child abuse. A 2018 report found that approximately 50% of children in Rwanda experience various forms of violence, including sexual, physical and emotional abuse [52]. Such experiences may shape caregiving behaviours later in life.

### Child age and the escalation of maternal disciplinary practices

Child age also appeared to influence disciplinary practices. Children aged ≥12 months were more likely to experience both physical and non-physical punishment compared to children younger than one year. Similar patterns have been observed in other settings. For example, studies conducted in the United States have reported increasing use of spanking as children grow older [49, 53]. As children enter toddlerhood, their growing independence and increased mobility can lead to behaviours that parents may find difficult to manage. This stage of development may prompt mothers to adopt stricter disciplinary measures. A study conducted in Tokyo found that mothers’ parenting stress at 1 and 36 months postpartum was a predictor of the use of physical punishment, even when controlling for maternal depressive symptoms [54].

### Maternal IPV and its influence on child discipline

Another important observation from this study is the association between women’s exposure to IPV and the use of harsher child disciplinary practices. Women who reported experiencing sexual, physical, or psychological IPV were more likely to report punitive behaviours towards their children. This finding aligns with previous research highlighting the interconnected nature of domestic violence and child maltreatment [55]. Even when children are not the direct targets of violence, exposure to household violence may affect their well-being and caregiving environment [56]. In some countries (e.g. Sweden), it is considered a crime to physically harm or abuse a partner in front of a child [57]. Women subjected to domestic violence frequently face challenges in their caregiving roles, which can result in suboptimal breastfeeding practices and inadequate nutrition for their children [58]. Furthermore, these women may exhibit increased aggression towards their children, including newborns [55]. The present findings also suggest that maternal exposure to multiple stressors particularly IPV and limited social support may create conditions in which harsh disciplinary practices are more likely. Women experiencing IPV often report higher emotional distress, and when this is combined with limited support from family or community networks, their coping capacity may be reduced [59]. These stressors do not operate in isolation; rather, they reinforce each other, creating a high-risk environment for the child. For example, IPV may undermine mothers’ mental health, reduce patience and tolerance during child-rearing, and limit a mother’s capacity to engage in responsive caregiving [60].

### Influence of mothers’ social support on child disciplining

The absence of social support further deprives the mother of practical help, advice, and emotional buffering, leaving her more likely to resort to punitive discipline [61]. This combination of mothers’ psychological strain, harsh disciplining behaviours, and inconsistent caregiving can directly and indirectly affect child nutrition through reduced breastfeeding frequency, irregular feeding, and heightened child stress ultimately increasing the risk of stunting [60]. Recognizing these interaction effects is essential for designing interventions that address both the psychosocial and practical needs of mothers, thereby fostering healthier growth and development in their children. Findings from our study have indicated that mothers who used punitive disciplinary measures were more likely to experience a lack of social support.

Consistent with prior studies, mothers reporting limited social support were more likely to report punitive disciplinary behaviours [62]. Furthermore, children of mothers who reported the use of physical punishment were more likely to be stunted. These results are consistent with research suggesting that early-life stressors, including child maltreatment or neglect, may be linked to impaired growth outcomes [63]. However, given the cross-sectional design of this study, these associations should not be interpreted as causal.

### Disrupted caregiving and early childhood stunting

Previous literature has proposed several mechanisms to explain how adverse caregiving environments may be associated with impaired child growth.

Child abuse and neglect have profound and lasting effects on children’s physical development contributing to stunting. Poverty and household conflicts contribute to increased mental health disturbances in mothers, which in turn affect their parenting practices [64]. These mothers may resort to a combination of physical and non-physical punishment for children as young as 1 month to 3 years. Such abusive disciplinary methods can have significant consequences on the child’s overall well-being. One significant impact of this behaviour is the disruption of breastfeeding. The mother’s emotional distress and use of punishment reduce both the quality and quantity of breastfeeding, leading to skipped meals and insufficient nourishment for the child. This issue is exacerbated when the child becomes lethargic from prolonged crying [65]. In addition, some mothers may even deprive their older children, those >1 year, of adequate feeding due to the emotional strain they are under [66]. This inconsistency in providing vital nutrition is a significant contributing factor to child stunting, particularly in the Northern Province of Rwanda. The lack of proper care, combined with the psychological and physical stress the mother faces, creates a cycle of malnutrition that negatively affects the child’s growth and development [66]. Addressing these underlying socio-economic and psychological challenges is crucial for tackling the root causes of stunting in this region. Our findings are consistent with previous research demonstrating that physical abuse or neglect in early childhood is associated with poor height-for-age outcomes, emphasizing how both direct and indirect stressors can have an impact on long-term growth trajectories in children [67, 68].

### Policy and Public Health Implications

The findings of this study have direct implications for child protection, mothers’ support, and nutrition policies in Rwanda, particularly for children ≤3 years of age. Rwanda has made significant progress in reducing stunting, yet our results highlight the urgent need to integrate parenting support and violence-prevention strategies into existing maternal and child health programs. Early parenting education delivered through community health workers and health facility-based antenatal and postnatal services should emphasize non-violent disciplining approaches and promote positive caregiving practices. Routine screening for intimate partner violence within maternal health services, coupled with timely referral to support services such as Isange One stop centers, can reduce the risk of intergenerational transmission of violence.

Strengthening community-based social support networks, such as “Umugoroba w’Ababyeyi” (Parents’ evening forum) and village-level solidarity groups, can help buffer the impact of maternal stress on child care. Finally, integrating psychosocial support and nutrition counselling into existing child growth monitoring and promotion programs will help address both the emotional and physical needs of mothers, thereby improving feeding practices and reducing stunting risk. These policy measures, when implemented together, could have a substantial impact on breaking the cycle of violence, poor caregiving, and child stunting in Rwanda.

### Methodological considerations

This study has notable strengths as well as limitations. It is among the first to examine the associations between mothers’ use of physical and non-physical punishment in children and child stunting in Rwanda, addressing a crucial gap in the scientific literature on early child development in low-resource settings. The study’s use of a representative sample enables insights into population-wide trends, enhancing the generalizability of its findings within rural Rwanda. However, several methodological limitations should be considered. The cross-sectional design limits the ability to infer causality because data collected at a single point in time cannot establish the direction of relationships between child abuse and stunting. A longitudinal study in the future could provide more robust evidence of causality by tracking changes over time and assessing the direction of these relationships. Another key limitation relates to the reliance on self-reported data for sensitive behaviours such as child disciplining methods and intimate partner violence. Social desirability bias may have led to underreporting of harsh disciplinary practices, as mothers might be reluctant to disclose behaviours perceived negatively. Similarly, self-reported IPV data may underestimate the true prevalence due to stigma, fear of reprisal, or cultural norms that discourage disclosure. These measurement biases could affect the observed associations, potentially attenuating the strength of the relationships. Additionally, we recognize that interviewer gender can influence responses in sensitive surveys; this has been noted as a limitation, and future studies could explore potential interviewer effects more systematically.

The study is based in Rwanda, therefore cultural and social factors specific to this context may limit the generalizability of the findings to other countries.

## Conclusion

This study found that children exposed to both physical and non-physical disciplining practices had nearly twice the odds of being stunted, even after accounting for maternal sociodemographic factors and IPV exposure. In addition, mothers exposed to IPV were more likely to use physical punishment on their children, further exacerbating child stunting. These findings highlight the urgent need for comprehensive interventions focusing on early childhood development, positive parenting strategies and widespread awareness of child protection laws throughout Rwandan communities.

To effectively address this issue, we propose integrating child abuse screening into paediatric clinical care protocols at all levels of child healthcare. Community health workers, healthcare providers and staff at Isange One Stop Centers (Isange One Stop Centers are integrated facilities established in Rwanda that provide comprehensive medical, psychosocial, legal, and police services to survivors of gender-based violence and child abuse under one roof) should be trained to recognize the signs and symptoms of child abuse and to understand the laws and policies against child abuse in Rwanda. Furthermore, as part of antenatal care, healthcare providers should educate expectant parents about the risks associated with any form of violence against children, regardless of age. In addition, nationwide awareness campaigns should be launched in community and school settings to raise awareness of the harmful effects of child abuse and promote a safer, healthier environment for children. By implementing these measures, Rwanda can strengthen its child protection and nutrition frameworks, and these findings may also inform similar strategies across other low-income countries where psychosocial stressors remain under-addressed in malnutrition frameworks.

## Supporting information

S1 File. Stata do file instructions. (PDF)

S1 Dataset. Minimal data exported to Stata.(CSV) S2 File. Questionnaire. (PDF)

## Data Availability

All relevant data are within the manuscript and its Supporting Information files

## Acknowledgements

This research was conducted as part of the Rwanda-Sweden partnership program, supported by the Swedish International Development Cooperation Agency (SIDA). We deeply appreciate the women and children who took part in the study and shared their personal experiences.

Special thanks are extended to the University of Rwanda and the five Swedish universities (University of Gothenburg, Umeå University, Lund University, Uppsala University and the Swedish University of Agricultural Sciences) for their essential logistical and supervisory assistance throughout the study.

